# Self-reported decreases in the purchases of selected unhealthy foods resulting from the implementation of warning labels in Mexican youth and adult population

**DOI:** 10.1101/2023.11.22.23298843

**Authors:** Alejandra Contreras-Manzano, Christine M. White, Claudia Nieto, Kathia L. Quevedo, Jorge Vargas-Meza, David Hammond, James F. Thrasher, Simón Barquera, Alejandra Jáuregui

**Affiliations:** Center for Nutrition and Health Research, National Institute of Public Health, Cuernavaca, Mexico; National Council for Humanities, Science and Technology, Mexico City, Mexico; School of Public Health Sciences, University of Waterloo, Waterloo, Canada; El Poder del Consumidor A.C. Mexico City, Mexico; Department of Health Promotion, Education and Behavior, Arnold School of Public Health, University of South Carolina, Columbia, SC, USA

**Keywords:** warning labels, food purchasing behavior, Nutrition labeling, Food policy, sugar sweetened beverages

## Abstract

**Background:** Front-of-package nutritional warning labels (WLs) are designed to facilitate identification and selection of healthier food choices. We assessed self-reported changes in purchasing different types of unhealthy foods due to WLs in Mexico and the association between the self-reported reductions in purchases of sugary beverages and intake of water and sugar-sweetened beverages.

**Methods:** Data came from 14-17 year old youth (n=1,696) and adults ≥18 (n=7,775) who participated in the Mexican arm of the 2020-2021 International Food Policy Study, an annual repeat cross-sectional online survey. Participants self-reported whether the WLs had influenced them to purchase less of each of ten unhealthy food categories due to WLs. Among adults, a 23-item Beverage Frequency Questionnaire was used derive past 7-day intake of water and sugary beverages analyzed to determine the relationship between self-reported reductions in purchasing sugary drinks due to the WLs. Multilevel mixed-effects logistic regression models were fitted to estimate the percentage of participants who self-reported reducing purchases within each food group, and overall. Sociodemographic characteristics associated with this reduction were investigated as well.

**Results:** Overall, 44.8% of adults and 38.7% of youth reported buying less of unhealthy food categories due to the implementation of WL, with the largest proportion reporting decreased purchases of cola, regular and diet soda. A greater impact of WLs on the reported purchase of unhealthy foods was observed among the following socio-demographic characteristics: females, individuals who self-identified as indigenous, those who were overweight, individuals with lower educational levels, those with higher nutrition knowledge, households with children, and those with a significant role in household food purchases. In addition, adults who reported higher water intake and lower consumption of sugary beverages were more likely to report reduced purchases of sugary drinks due to the WLs. Adults who reported greater water intake and lower sugary beverages intake were significantly more likely to report buying fewer sugary drinks due to the WLs.

**Conclusion:** Our findings suggest that implementation of WLs has reduced purchases of unhealthy foods in Mexico. These results underscore the positive impact of the labeling policy particularly in subpopulations with lower levels of education and among indigenous adults.

## Background

Front-of-pack nutrition labeling (FoPL) provides simple, easy to find information about nutrients to promote healthier dietary habits and, thereby, reduce diet-related non-communicable chronic diseases [1,2]. Various FoPL systems have been introduced in different countries, aiming to enhance consumer decision-making capacity when comparing and purchasing foods with better nutritional profiles [3–6]. These FoPL systems include warning labels (WLs), which identify products that contain excessive amounts of nutrients that may be harmful to health [7, 8]. However, given that WLs have only been recently implemented, to date, scarce evidence exists on their real-life impacts.

In Mexico, WLs were implemented in October 2020 as a public health policy [9] in response to the increasing demand for processed and ultra-processed food products, including sugary drinks, and the prevalence of overweight, obesity, diabetes, and other diet-related non-communicable diseases in the country, with important economic and health repercussions [10]. Mexican WLs consist of black octagon-shaped warnings prominently displayed on the front of packaged foods to help people identify products that have “excess” levels of five nutrients of concern: calories, added sugars, saturated fat, trans fat, and sodium [9]. As part of the policy, two precautionary disclaimers are also displayed as black rectangles on the front of packages are also included to discourage child consumption of products with caffeine (‘Contains caffeine, avoid in children’) or non-sugar sweeteners (‘Contains sweeteners, not recommended in children’) [9]. Experimental and qualitative studies conducted with various FoPL systems consistently demonstrate that WLs are the best understood by Mexican consumers from different backgrounds, including low education and low-income groups [11–14]

Processed and ultra-processed products, such as sugary drinks, fruit nectars, savory snacks, and cakes, have a high density of calories and nutrients of concern (i.e., saturated fat, trans fat, sugar, sodium) linked with the development of non-communicable diseases [15]. However, consumers in general are often unaware of the risks and consequences of choosing products with poor nutritional quality; consequently, decisions are usually made based on taste, smell, texture, and/or marketing [15]. A review of experimental studies found that WLs may discourage consumers from purchasing products with excess levels of critical nutrients, including sugary drinks [17].

Few ‘real-world’ studies have evaluated the impact of WLs or other FoPLs on the purchasing behavior of consumers [18,19]. A study of Chilean households reported that after the introduction of black octagon WLs, among purchases of products with WLs, overall calories, calories from sugar, calories from saturated fat and sodium purchased decreased by −49.4 kcal, −20.7 kcal, −6.2 kcal and −96.6 mg per capita/day respectively. Larger absolute reductions in sugar were found for beverages, whereas foods had larger absolute reductions in sodium and saturated fat [18]. Additionally, the volume of purchases of beverages with “high in” WLs in Chile decreased by 22.8 mL/capita/day post-regulation, when compared to the expected purchases based on pre-regulation trends [19]. For both, more highly educated households had a greater reduction in purchases of beverages with WLs compared to households with lower levels of education. These results are consistent with a study conducted among Chilean parents, where 49.5% reported having stopped buying certain foods due to the presence of a WLs [20]. A lower probability of food purchase pattern change was observed in families that did not consider the WLs an important tool, did not understand the WLs, and did not regularly read nutritional labels prior to law implementation [20].

Evaluations of the Chilean WLs policy suggest that it has reduced consumption of unhealthy food and that differential effects across population groups are important to assess. To date, no studies have evaluated changes in purchasing behavior following WLs implementation in Mexico. Therefore, this study aimed to analyze self-reported decreases in purchases of unhealthy foods due to WLs in Mexico, and secondly, to investigate associations between the self-reported reductions in purchasing sugary beverages and past 7-day consumption of water and sugar-sweetened beverages.

## Methods

### Study design

The current study used data from the Mexican arm of the 2020 and 2021 International Food Policy Study (IFPS). The IFPS is an annual repeated cross-sectional online survey of children (10-13 years), youth (14-17 years) and adults (≥18 years) from multiple countries, including Mexico. Detailed methods on the IFPS can be found elsewhere [21, 22]. Data were collected in November-December in both 2020 and 2021, one month and over one year after the after the enactment of the Mexican WLs, respectively. Surveys for all age groups asked participants to report the usefulness of WLs, with only those aged 14 or older asked whether the WLs had caused them to change their food purchasing behavior. Hence, we only analyze data from those aged 14 or older.

### Sample & Recruitment

Participants were primarily recruited through Nielsen’s Consumer Insights Global Panel and their partner panels. Additionally, an oversample of adult respondents with lower educational attainment recruited through Qualtrics and their partner panels in 2021. In prior waves, recruitment through Nielsen and their partners yielded a sample with higher educational attainment than observed in the general population, thus the oversample was recruited in 2021 to obtain a sample that more closely resembles the national education distribution. Participants were recruited using both probability and non-probability sampling methods. Eligible participants were adults 18 to 100 years of age and youth aged 14-17 years residing in Mexico.

For the adult survey, email invitations with unique survey access links were sent to a random sample of panelists in Mexico after targeting for demographics; panelists known to be ineligible were not invited. Potential respondents were screened for eligibility and quota requirements based on age and sex. In 2021, respondents recruited as part of the low education oversample also had to report having a high school-level education or less. After screening, all potential respondents were provided with information about the study and were asked to provide consent before participating.

For the youth survey, parents/ guardians were provided information about the study and asked for permission for their child to participate. Only one child per household was invited to participate. The child was subsequently screened directly to confirm eligibility based on age. Eligible children were provided with information about the study and were asked to provide consent before participating. The youth’s parent/ guardian and adult respondents received remuneration in accordance with their panel’s usual incentive structure (e.g., points-based, or monetary rewards that can be redeemed for e-gift cards, catalog items, cash, donations and/or chances to win monthly prizes).

The surveys were conducted in Spanish. The cooperation rate for adults was 9.2% and 16.1% for 2020 and 2021 and for youth 3.4% and 7.4% respectively. Participants who answered “don’t know” or “refuse to answer” in any of the measures of interest were excluded from final analyses. The final analytic sample with complete information for this study consisted of 7,775 adults (n=3,900 in 2020 and n=3,875 in 2021), and 1,696 youths aged 14 to 17 years (n=891 in 2020 and n=805 in 2021). The low-education level oversample added 2,072 adults to the main sample in 2021 resulting in an analytic sample of 9,847 adult participants for both years.

The study was reviewed by and received ethics clearance through a University of Waterloo Research Ethics Committee, the Institutional Review Board at the University of South Carolina (only 2021 survey), and the Research Ethics Committee at the Instituto Nacional de Salud Pública in Mexico.

### Self-reported changes in unhealthy food purchases

Participants reported changes in their food purchases of nine different food categories where WLs were common: cola (Coca-Cola, Pepsi, etc.), soda (Sprite, Orange Crush, etc.), diet soda (Coca-cola Zero, Pepsi de Dieta, etc.), sweetened fruit drinks (lemonade, iced tea, SunnyD, fruit punch/cocktail, etc.), candy or chocolate bars, snacks such as chips, desserts such as cakes, cookies and ice cream, and sugary cereals. Emerging research suggests that 100% fruit juice may similarly amplify the risk of mortality as observed for sugary drinks [23]. Although 100% fruit or vegetable juice is not included in the WLs regulation, fruit beverages are widely consumed in Mexico [24]. Therefore, self-reported changes in the purchases of processed 100% fruit or vegetable juice were also assessed with the intention of incorporating a “comparison group” without WLs in the analysis.

Participants were shown an image of the “excess calories” WLs and asked how often they had seen that type of food label on packages on in stores (Adults: 4,284 in 2020 and 4,172 in 2021. Youth: 904 in 2020 and 829 in 2021). On average, 98.5% of participants reported having ever seeing the WLs and those 10,043 youth and adults were asked for each of the food categories: “*Have the warning labels (black octagons) changed whether you buy the following packaged products for you or your family?”* The response options were: “Buy less”, “Buy more”, “No change”, “Don’t know” or “Refuse to answer”. Participants were classified as: 0 “No change/Buy more” or 1 “Buy less”.

### Self-reported usefulness of nutrient-specific warning labels

The self-reported usefulness of each nutrient-specific WLs was assessed by showing participants an image of each of the WLs and asking: *“Which of these stamps, if any, has been most useful to choose healthier foods?”* Respondents could select one of the following WL: “Excess Calories”, “Excess Sodium”, “Excess Saturated Fat”, “Excess Trans Fat”, “Excess Sugars”, or could select “None of the stamps have been useful”, “All of the stamps have been equally useful”, “Don’t know” or “Refuse to answer”.

### Beverage intake in adults

Beverage intake among adults was assessed through a Beverage Frequency Questionnaire (BFQ) that queried 23 beverage categories [25]. Participants were asked: *“During the past 7 days, how many drinks did you have in each category below?”* For each category of drink consumed, participants were shown common beverage container sizes and asked to select the usual size of drink consumed, which was subsequently used to estimate the total volume consumed in the last 7 days per beverage category. The intake of “sugar sweetened beverages” was estimated by summing the estimated volume of regular soda, sweetened fruit drinks, regular flavored waters or vitamin waters with calories, regular sports drinks, regular energy drinks, chocolate milk, coffee/tea with sugar, sweetened specialty coffees/teas, and sweetened smoothies/protein shakes/drinkable yogurt. Water intake was estimated by summing the estimated volume of tap and plain bottled water.

#### Covariates

Covariates assessed included age, sex at birth (male; female), indigeneity (self-identifying as indigenous or non-Indigenous), self-reported nutrition knowledge (responses recoded as not knowledgeable, somewhat knowledgeable, or knowledgeable). Education level for adults was based on the highest level of formal education completed and recoded as low=basic or less; medium=technical or commercial studies with completed secondary education or less; high= technical or commercial studies with completed high school education or higher.

Perceived income adequacy (*“Thinking about your total monthly income, how difficult or easy is it for you to make ends meet?”)* was assessed with a 5-point Likert scale with responses recoded as “difficult” (very difficult/difficult), “neither easy nor difficult”, or “easy” (easy/very easy). BMI was calculated using reported weight and height and categorized according to World Health Organization (WHO) standards for adults and z-scores for age and sex using a WHO macro for youth [26]. Food shopping role for adults was assessed with the question: *“How much of the food shopping do you do in your household?”* with the responses recoded as “Most” or “Some or None”.

#### Statistical analysis

Post-stratification sample weights for the main sample were constructed based on known population totals by age, sex at birth, region, and indigeneity; for the sample considering the oversample in 2021, sample weights additionally accounted for education. Estimates reported are weighted.

We used two modelling approaches to estimate the self-reported changes in food purchases across survey years and the associated factors with these changes. For all analyses, separate regression models were estimated for youth and adults. Firstly, to examine a general effect of the WLs in the self-reported changes in unhealthy purchases across various food groups (0= no change/buy more; 1= buy less), we employed a repeated measures (mixed effects) logistic regression model introducing multiple observations for each respondent corresponding the different types of food groups. This model did not include 100% fruit juice because this product does not have any WLs. Instead, we used diet soda as the reference group since it has fewer warning labels compared to the other food categories. The individual was considered as the clustering level. The models for youth and adults were adjusted by survey year, sex, age, indigeneity, income adequacy, and BMI category. For adults, the model was further adjusted by education level, children in the household, self-reported nutrition knowledge, and food shopping role. Additionally, we fitted a mixed-effects logistic regression model for self-reported reduction in purchasing sugary drinks, with quintiles of past 7-day water intake and of past 7-day sugary drink intake as the independent variable, same covariates described previously were used to adjust the model.

Secondly, we used separate logistic regression models for each of the ten food products, including 100% fruit juice, to estimate the correlates of self-reported reducing purchases, adjusting for the same covariates as the previous models.

To analyze differences in the self-reported usefulness of each nutrient-specific WLs we estimated separate logistic regression models for adults and youth.

Secondary analyses used the same statistical approach after including the 2021 oversample of participants with low education. The individual was considered as the clustering level. Statistical significance was set with a p value of 0.05. Analyzes were performed in Stata 14.0v.

## Results

Participants characteristics were somewhat consistent across study years. The mean age for youth was 15.4 years and 40.7 years for adults. The male proportion was 50.6% for youth and 48.1% for adults; indigeneity was 22.6% in youth and 19.2% adults; the percentage of participants with high income adequacy was 20.8% in youth and 16.5% in adults. Around 5% of youth and 15% of adults had a BMI ≥30. Half of adults had children in their household, 73.4% had an important food shopping role in the household. However, differences between study years were observed in the distribution of income adequacy, BMI, education level, and nutrition knowledge categories among adults. (Table 1) The analysis considering the oversample of individuals with low education in 2021 had a higher percentage of participants with a low education level (74.6%), or an important role in food shopping within the household (68.2%), and a lower percentage with perceived nutrition knowledge (9.4%) or a BMI <25 (32.6%), compared to the main survey sample for 2021 (Additional File 1).

**Table 1.** Sociodemographic characteristics of youth and adult participants, International Food Policy Study, 2020 and 2021.

| n sample |  | Youth (13-17 y) |  |  | Adults (<sup>a</sup>18 y) |  |  |
| --- | --- | --- | --- | --- | --- | --- | --- |
|  |  | 2020 | 2021 | Both | 2020 | 2021 | Both |
|  |  | 891<br>% (95% CI) | 805<br>% (95% CI) | 1,696<br>% (95% CI) | 3,900<br>% (95% CI) | 3,875<br>% (95% CI) | 7,775<br>% (95% CI) |
| Age (years)* |  | **15.3 (15.2, 15.4)** | **15.5 (15.4, 15.6)** | 15.4 (15.3, 15.4) | 40.5 (39.9, 41.1) | 40.9 (40.3, 41.6) | 40.7 (40.3, 41.2) |
| Sex | Female | 49.4 (45.4, 53.4) | 49.3 (45.1, 53.5) | 49.4 (46.5, 52.2) | 51.8 (49.9, 53.7) | 52.0 (50.2, 53.8) | 51.9 (50.6, 53.2) |
|  | Male | 50.6 (46.6, 54.6) | 50.7 (46.5, 54.9) | 50.6 (47.7, 53.5) | 48.2 (46.3, 50.0) | 47.9 (46.1, 49.8) | 48.1 (46.7, 49.4) |
| Indigeneity | Yes | 20.9 (17.1, 25.2) | 24.6 (20.5, 29.2) | 22.6 (19.8, 25.7) | 19.2 (17.6, 20.9) | 19.2 (17.7, 20.9) | 19.2 (18.1, 20.4) |
|  | No | 79.1 (74.8, 82.9) | 75.4 (70.8, 79.5) | 77.4 (74.3, 80.2) | 80.8 (79.1, 82.4) | 80.8 (79.1, 82.3) | 80.7 (79.6, 81.9) |
| Income adequacy | Easy | 38.9 (34.9, 43.0) | 21.5 (18.4, 25.1) | 20.8 (18.6, 23.2) | 13.7 (12.6, 15.0) | **19.4 (18.0, 20.8)** | 16.5 (15.6, 17.5) |
|  | Neither | 38.6 (34.9, 42.5) | 38.9 (34.9, 43.0) | 38.8 (36.0, 41.6) | 36.1 (34.3, 37.9) | **40.8 (39.0, 42.7)** | 38.4 (37.2, 39.7) |
|  | Difficult | 41.2 (37.2, 45.3) | 39.5 (35.4, 43.7) | 40.4 (37.6, 43.4) | 50.2 (48.3, 52.1) | **39.8 (37.9, 41.7)** | 44.9 (43.6, 46.3) |
| BMI category | <25 | 53.5 (49.5, 57.5) | 55.0 (50.8, 59.2) | 54.2 (51.3, 57.1) | 38.0 (36.2, 39.8) | **41.1 (39.3, 43.0)** | 39.6 (38.3, 40.8) |
|  | 25-29 | 22.1 (19.0, 25.5) | 19.1 (16.1, 22.6) | 20.7 (18.5, 23.1) | 31.9 (30.2, 33.8) | **30.6 (28.9, 32.3)** | 31.2 (30.0, 32.5) |
|  | ≥30 | 4.4 (3.1, 6.2) | 4.9 (3.5, 6.9) | 4.6 (3.6, 5.9) | 15.3 (13.9, 16.8) | **14.8 (13.5, 16.2)** | 15.1 (14.1, 16.1) |
|  | Missing | 20.0 (16.9, 23.6) | 20.9 (17.7, 24.4) | 20.5 (18.2, 22.9) | 14.7 (13.4, 16.1) | **13.4 (12.1, 14.7)** | 14.0 (13.1, 15.0) |
| Education level | Low |  |  |  | 21.4 (19.9, 23.0) | **19.1 (17.7, 20.5)** | 20.2 (19.2, 21.3) |
|  | Medium |  |  |  | 13.6 (12.2, 14.9) | **14.2 (12.9, 15.6)** | 13.8 (12.9, 14.8) |
|  | High |  |  |  | 65.0 (63.2, 66.8) | **66.8 (64.9, 68.5)** | 65.9 (64.6, 67.1) |
| Children in the household (<18 y old) | No |  |  |  | 51.2 (49.3, 53.1) | 50.4 (48.5, 52.3) | 50.8 (49.5, 52.1) |
|  | Yes |  |  |  | 48.7 (46.8, 50.6) | 49.6 (47.7, 51.4) | 49.2 (47.8, 50.5) |
| Food shopping role (resume) | Most |  |  |  | 73.6 (72.0, 75.2) | 73.2 (71.6, 74.8) | 73.4 (72.2, 74.6) |
|  | Some or None |  |  |  | 26.4 (24.7, 28.0) | 26.7 (25.1, 28.4) | 26.5 (25.4, 27.7) |
| Nutrition knowledge | Not knowledgeable |  |  |  | 31.4 (29.7, 33.1) | **33.9 (32.2, 35.7)** | 32.7 (31.4, 33.9) |
|  | Somewhat |  |  |  | 55.5 (53.6, 57.4) | **50.8 (48.9, 52.7)** | 53.2 (51.8, 54.5) |
|  | Knowledgeable |  |  |  | 13.1 (11.8, 14.5) | **15.2 (13.9, 16.6)** | 14.2 (13.2, 15.1) |

\*Means. **Bold numbers** indicate p value <0.05 among study years.

Figure 1 shows the adjusted percentage of adults and youth reporting that the WLs had led them to buy less of each of the studied food categories and overall. On average, 44.8% of adults and 38.7% of youth self-reported that the WLs had led them to buy less of the food categories overall. Sugary beverages, such as cola, soda, diet soda, and sweetened fruit drinks had the largest self-reported decreases among adults (50.4-52.4%) and youth (41.0-49.9%). Meanwhile, 100% fruit juice, a product free of WL, had the smallest self-reported decreases among youth (19.9%) and adults (28.1%). Additional file 2 shows the adjusted percentage of participants who self-reported buying less of each food group and overall, in adults and youth by study year. No differences were observed between 2020 and 2021 in the percentage of participants reporting buying less food products overall and across food categories; except for 100% fruit juice, where the decrease in purchases was lower in 2021 (26.7%) compared to 2020 (29.6%, p<0.05). Secondary analyses including the oversample of low education adults in 2021 showed a slightly higher percentage of participants reporting that the WLs had led them to buy less unhealthy food products overall (48.8%) in comparison to the main sample, and a higher percentage of participants reporting that the WLs had led them to buy less candy or chocolate bars in 2021 (50.9%) compared to 2020 (47.6%) (Additional file 2).

**Figure 1.**
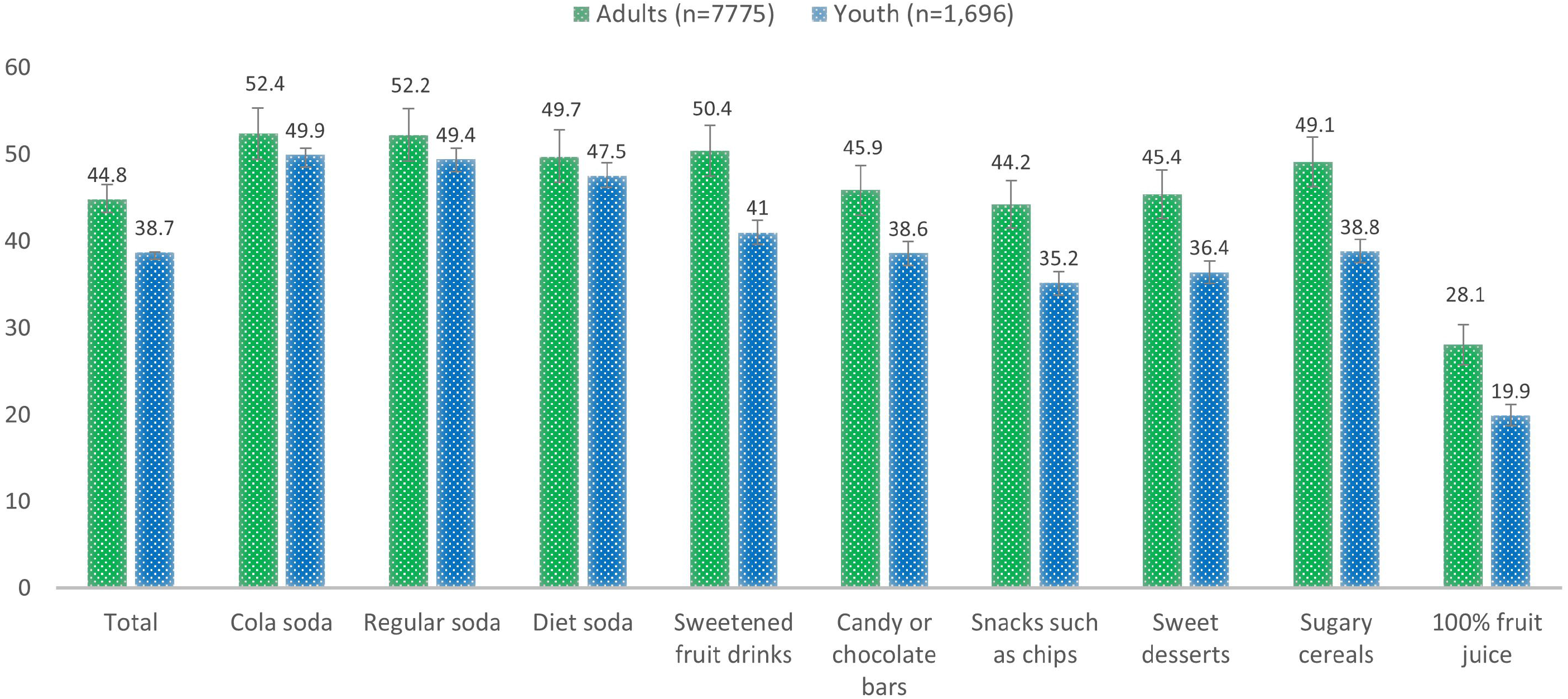
Adjusted percentage (%) of Mexican adults and youth reporting decreases in food purchases of selected food categories after the enactment of warning labels, International Food Policy Study, 2020 and 2021. Percentage of participants reporting decreases in food purchases derived from logistic regression models adjusted for age, sex, indigeneity, income adequacy and BMI category for adults and youth, and additionally adjusted by educational level, children in the household, nutrition knowledge, role in the food shopping in the household for adults. The overall percentage was derived from a multilevel logistic regression model adjusted by the same covariates and food group categories (diet soda as the reference); individuals were considered the clustering level. The multilevel model did not include 100% fruit juice because this product does not have any warning labels. All percentages were estimated using margins.

Figure 2 shows the odds ratio of reporting that the WLs had led them to purchase fewer sugary drinks across quintiles of sugary beverage or water intake among adults. Participants classified in the highest quintile of intake of sugary drinks had lower odds of reporting to buy less sugary drinks (OR: 0.09, p<0.05) compared to those classified in the lowest quintile. Conversely, those classified in the highest quintile of water consumption had higher odds of reporting to buy less sugary drinks (OR: 4.80, p<0.05) compared to those in the lowest quintile of water consumption. The analysis of the oversample with a low education level yielded similar results (Additional Figure 1).

**Figure 2.**
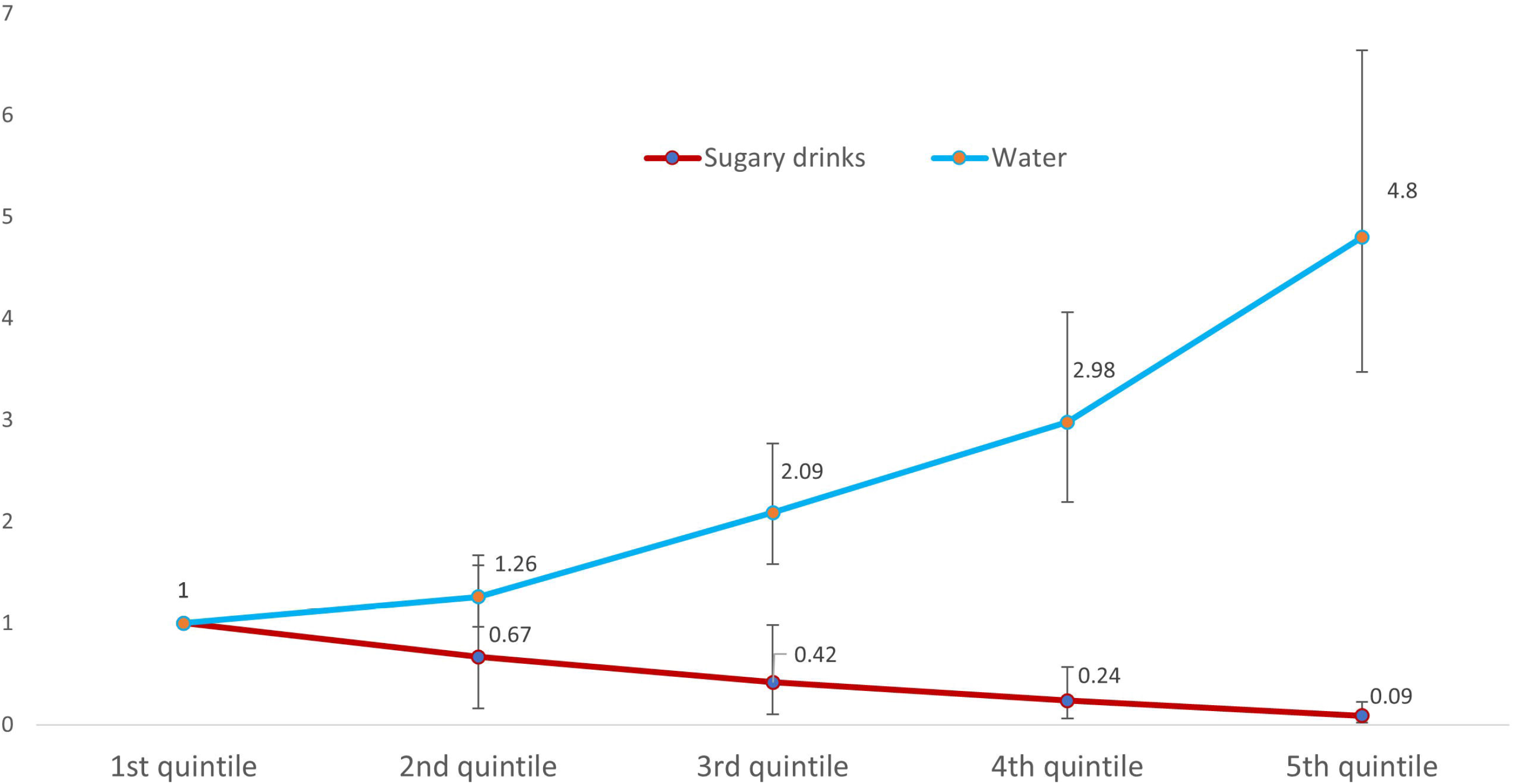
Odds ratio of self-reported buying less sugary drinks due to the WLs by quintile of past 7 day intake of sugary beverages and water in adults, International Food Policy Study, 2020 and 2021 (n=7,071). Self-reported purchase change in sugary drink groups included cola, soda, and sweetened fruit drinks. Sugary drink intake was based on the summed volume of regular soda, sweetened fruit drinks, regular flavored waters or vitamin waters with calories, regular sports drinks, and regular energy drinks. Median intake per quintile: Sugary drinks intake (Q1=71.4 ml, Q2=301.7 ml, Q3=540 ml, Q4=848 ml, Q5=1528 ml), Water intake (Q1=0 ml, Q2=357 ml, Q3=750 ml, Q4=1071 ml, Q5=1857 ml). Multilevel regression model adjusted by age, sex, indigeneity, educational level, income adequacy, children in the household, nutrition knowledge, role in the food shopping in the household, BMI category and year of the survey.

Table 2 shows the sociodemographic characteristics associated with self-reported changes in food purchases due to the WLs. Among adults, reporting that the WLs had led them to buy less unhealthy food products was positively associated with being female, older age, self-identifying as indigenous, having a BMI between 25 and 29, low educational level (compared to high), having children in the household, self-reporting higher nutrition knowledge, and having an important role shopping food in the household. Among youth, perceiving to buy fewer food products due to the implementation of the WLs was negatively associated with age and positively associated with indigeneity and low-income adequacy.

**Table 2.** Adjusted odds ratios for perceiving buying less unhealthy foods due to the warning labels, International Food Policy Study, 2020 and 2021.

|  |  | Youth (n=1,678) |  | Adults (n=7,775) |  |
| --- | --- | --- | --- | --- | --- |
|  |  | OR (95% CI) | p value | OR (95% CI) | p value |
| **Survey year** | 2020 | 1.0 | Ref. | 1.0 | Ref. |
|  | 2021 | 0.83 (0.62, 1.12) | 0.219 | 0.9 (0.78, 1.04) | 0.143 |
| **Sex** | Male | 1.0 | Ref. | 1.0 | Ref. |
|  | Female | 1.27 (0.94, 1.72) | 0.117 | 1.59 (1.37, 1.83) | <0.001 |
| **Food group** | Diet soda | 1.0 | Ref. | 1.0 | Ref. |
|  | Cola | 1.33 (1.03, 1.72) | 0.031 | 1.33 (1.19, 1.49) | <0.001 |
|  | Soda | 1.27 (0.99, 1.62) | 0.057 | 1.3 (1.17, 1.44) | <0.001 |
|  | Sweetened fruit drinks | 0.53 (0.4, 0.71) | <0.001 | 1.07 (0.95, 1.21) | 0.237 |
|  | Candy or chocolate bars | 0.43 (0.32, 0.58) | <0.001 | 0.67 (0.58, 0.77) | <0.001 |
|  | Snacks | 0.3 (0.22, 0.4) | <0.001 | 0.56 (0.49, 0.64) | <0.001 |
|  | Dessert | 0.34 (0.26, 0.46) | <0.001 | 0.64 (0.56, 0.73) | <0.001 |
|  | Sugary cereals | 0.44 (0.32, 0.58) | <0.001 | 0.94 (0.82, 1.07) | 0.34 |
| **Age** |  | 0.86 (0.75, 0.98) | 0.024 | 1.01 (1, 1.01) | 0.024 |
| **Indigeneity** | Non-indigenous | 1.0 |  | 1.0 | Ref. |
|  | Indigenous | 2.47 (1.61, 3.79) | <0.001 | 3.15 (2.59, 3.82) | <0.001 |
| **Income adequacy** | Easy | 1.0 | Ref. | 1.0 | Ref. |
|  | Neither | 1.32 (0.91, 1.91) | 0.140 | 0.81 (0.66, 0.99) | 0.041 |
|  | Difficult | 1.59 (1.08, 2.34) | 0.017 | 0.87 (0.71, 1.07) | 0.199 |
| **BMI category** | <25 | 1.0 | Ref. | 1.0 | Ref. |
|  | 25-29 | 0.75 (0.52, 1.09) | 0.130 | 1.35 (1.14, 1.6) | 0.001 |
|  | ≥30 | 0.6 (0.31, 1.17) | 0.134 | 1.03 (0.83, 1.28) | 0.780 |
|  | Missing | 1.08 (0.73, 1.6) | 0.688 | 1.43 (1.14, 1.78) | 0.002 |
| **Educational level** | High |  |  | 1.0 | Ref. |
|  | Medium |  |  | 1.08 (0.88, 1.34) | 0.453 |
|  | Low |  |  | 1.64 (1.37, 1.98) | <0.001 |
| **Children in the household (<18 y old)** | No |  |  | 1.0 | Ref. |
|  | Yes |  |  | 1.42 (1.23, 1.64) | <0.001 |
| **Nutrition Knowledge** | Not knowledgeable |  |  |  |  |
|  | Somewhat know |  |  | 2.09 (1.79, 2.45) | <0.001 |
|  | Knowledgeable |  |  | 2.89 (2.29, 3.64) | <0.001 |
| **Shopping role** | Some or none |  |  |  |  |
|  | Most |  |  | 1.93 (1.64, 2.28) | <0.001 |

Multilevel mixed-effects logistic regression adjusted by food group and the covariates shown in the table. \*All food categories were included except 100% fruit or vegetable juice.

Figure 3 shows the adjusted self-reported usefulness of each nutrient-specific WLs among youth and adults in both survey years. On average, 31.3% (95% CI: 28.6-33.9) of youth and 32.1% (95%CI: 30.9-33.4) of adults self-reported that all the nutrient-specific WLs were equally useful for choosing healthier foods, with the next highest percentage reporting that the “Excess Sugars” warning was most useful (23.3% and 18.2% respectively). Around 15% of youth (95% CI: 13.2-17.4) and adults (95% CI: 15.7-17.7) self-reported that none of the nutrient-specific WLs had been useful. The “Excess sodium” label and the “Excess Trans fat” label were rated most useful by the lowest proportion of youth (4.3%), and adults (6.3%), respectively. These percentages remained similar across study years, except for “All have been equally useful” which decreased from 34.1% in 2020 to 29.6% in 2021 (p<0.05) (Additional file 3). Some factors associated with the self-reported usefulness of some WLs among adults were; age with “Excess Calories”, “Excess Sugars”, “None” and “All”, educational level for “Excess calories” and “All”, indigeneity for “Excess calories”, “Excess Sugars” and “None”, and having children in the households for “Excess Sodium”, “Excess Sugars”, “None” and “All” (Additional file 4).

**Figure 3.**
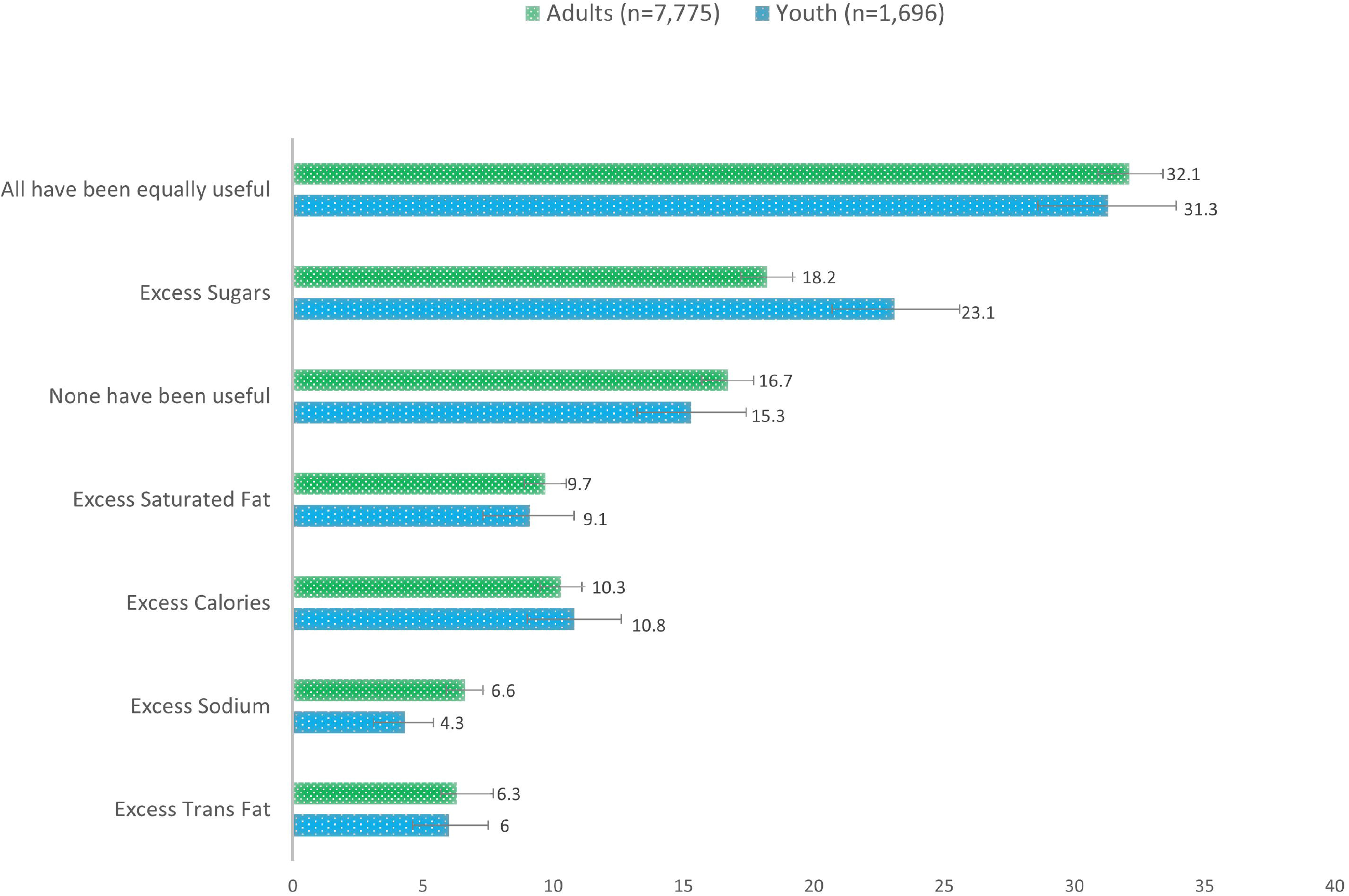
Adjusted percentage (%) of self-reported usefulness of each nutrient-specific warning labels among Mexican adults and youth. International Food Policy Study, 2020 and 2021. Data derived from a logistic regression model adjusted by age, sex, indigeneity, income adequacy and BMI category for youth and adults, and additionally adjusted by educational level, children in the household, nutrition knowledge, role in the food shopping in the household for adults.

## Discussion

The results of this study show that, one month and one year after the enactment of WLs in Mexico, more than a third of youth and nearly half of adults self-reported that this labeling system had led them to decrease the purchase of various unhealthy food products. Self-reported decreases in purchases of unhealthy food products were associated with various demographic characteristics, including indigeneity, having a lower education (adults), or lower income adequacy (youth). Larger decreases in self-reported purchases of sugary drinks were observed among those with the highest water consumption but not among those with high consumption of sugary drinks. These findings demonstrate a positive impact of the labeling policy, which aims to promote healthier food choices.

Scarce evidence exists regarding the real-life effect of FoPLs on food purchases. In Chile, a pioneering country in the implementation of WLs in 2016, comparisons before and after the implementation of this labeling have shown decreases in the overall purchases of calories, sugar, saturated fat, and sodium in the school setting [18], as well as in the volume of high-sugar beverage purchases [19]. This study adds to the literature by investigating the self-reported effect of WLs on unhealthy food purchases following the implementation of WLs in Mexico, finding that WLs may have led consumers to decrease their purchases of unhealthy food categories with sustained perceived effects at least one year after the introduction of the labels. These results are in line with recent meta-analyses suggesting that food labeling reduces consumer intake of energy, total fat, and other unhealthy dietary options, while increasing the selection of healthier products and vegetable consumption [27, 28]. Similarly, previous research has also shown that following the implementation of WLs in Mexico, increases in the awareness, use and understanding of nutrition labels were observed among Mexican adults and youth [29, 30]. Additionally, our study found that only 15% of the surveyed adults and young individuals considered none of the labels to be useful, indicating that the majority of the population perceives WLs as a useful tool for decision-making.

Data from the National Health and Nutrition Survey (ENSANUT) of Mexico [31] showed decreases in the percentage of Mexican adults reporting daily consumption of sugary beverages between 2020 (86.7%) and 2021 (69.3%) [32]. According to our study results, the impact of the implementation of WLs in Mexico might have contributed to these declines.

Our findings show that the self-reported impact of WLs was similar between 2020 and 2021 with a slightly higher prevalence of reduced purchases in 2021. This difference may be attributable to the timing of our study. Towards the end of 2020, WLs were newly introduced [34], and not all products were properly labeled, and some items might still have retained the previous labeling system. However, by the end of 2021, the WLs were fully implemented, facilitating their accurate utilization by consumers, and possibly showing a wear-in period of label influence as more products use them and consumers get accustomed to using WLs [33]. Future research should assess the continued impact of the labels over the long term to explore possible wear-out effects of the labels, as has been found for cigarette warning labels, where wear out also varies across sociodemographic groups of people who smoke [35].

In our study, we found that the largest self-reported decreases in unhealthy food purchases among both adults and young people occurred among sugary drinks, with lower self-reported reductions in other food categories such as desserts, snacks, or candies. While some research has suggested that WLs may have more impact in reducing the intake of sugar-sweetened beverages compared to other food categories [19], other research among Mexican children and adults suggests that the objective understanding of WLs is similar across food categories [36]. Indeed, this larger self-reported effect on sugary drinks may be attributable to Mexico being among the leading consumers of sugar-sweetened beverages globally [37] and sugary drinks being more commonly consumed than the other unhealthy products assessed [32]. According to the ENSANUT 2020 [29], sugary drinks were consumed daily by 90.9% of children aged 5-11 years, 90.7% of adolescents, and 86.7% of adults. Conversely, snacks, desserts, and sugary cereals were only consumed daily by less than 55% of the population. Furthermore, in addition to the traditional WLs indicating excess content of critical nutrients, many sugary drinks in Mexico are also required to display the two rectangle-shaped warning legends related to sweeteners and caffeine content [38]. These additional labels, which are generally less applicable to other food categories, may contribute to changes in the perceived healthiness of cola beverages and artificially sweetened drinks [38]. In line with the former, data from ENSANUT 2021 suggested that one year after the enactment of WLs in Mexico, 21.6% of adults identified excess calories in cola beverages, 82.3% identified excess sugars, 22.4% identified the caffeine warning, and 11.8% identified the sweeteners warning [32]. However, our results also indicate that self-reported decreases in the purchases of sugary drinks were more common among individuals who were in the highest quintile of water consumption, rather than sugary drinks. These findings underscore the importance of directing communication efforts to heavy consumers of sugary drinks to promote changes in the social norms regarding soda consumption in Mexico and complement current front-of-pack labeling and soda tax regulations in the region as well as additional policies aimed at improving the food environment [32].

In our study, participants with lower educational level, lower income adequacy and indigeneity status self-reported a greater reduction in the purchase of unhealthy foods because of the WLs. These population groups have a disproportionate burden of non-communicable diseases and lower access to healthy and nutritious foods [39]. Results of this study suggest that WLs may aid in decreasing health disparities. Indeed, WLs have been proposed as a policy with equitable impacts among consumers of all socioeconomic strata [12, 13, 36] that can have significant impacts even on populations with social inequities, who potentially may benefit more from public strategies that enhance food-related decision-making [27, 28]. Consistent with our findings, other studies have demonstrated that WLs can have a greater influence on food purchasing choices among specific subpopulations [11–13]. Experimental studies have shown that WLs are equally understood by consumers of all backgrounds and promote healthier purchasing intentions [11–13, 17–19, 32, 36].

WLs also led to greater self-reported decreases of unhealthy food purchases among adult women, those with children in the household, and those with overweight. Results suggests that in Mexico women may be more aware and willing to change purchasing decisions due to front-of-pack labeling than men. This is consistent with nationally representative studies showing that Mexican women were more likely to correctly interpret front-of-pack labels [41].

In line with our results regarding the association between BMI category and WLs, Sagaceta et al. (2019) showed that the WLs help Mexicans with overweight and obesity, as well as other non-communicable chronic diseases to classify unhealthy foods products more appropriately compared with GDA [42]. Our results are also in line with reports showing that the presence of WLs on food packages was associated with lower perceptions of healthfulness and purchasing intentions of Chilean parents [20, 43].

Finally, in our study, a greater proportion of participants rated the calorie, sugar, or saturated fat WLs as most useful, with the lowest proportion of adults rating the trans-fat WLs as most useful. This could be attributed to the fact that only around 1% of Mexican products are reportedly high in trans-fat, while 48.3, 40.4 and 33.9% of products were reportedly high in calories, sugar, and saturated fat, respectively, and showed the corresponding WLs [38]. Additionally, in Mexico, a law prohibiting the use of trans fats was approved in 2023, which is expected to lead to a decrease in the visibility and utilization of this label among the Mexican population [44].

This is the first study to describe the self-reported changes in unhealthy food purchases due to the implementation of FoPL in Mexico, contributing to the current knowledge on the impacts of front-of-pack labels at the population level. Self-reported changes in food purchasing have been used in other studies to evaluate the performance of nutrition policies [45–47] because they provide insight into individuals’ own perceptions and behaviors offering a unique perspective on how WLs may have impacted their daily lives and can capture nuances that objective measures might miss, such as emotional or psychological responses to interventions. The measure used to assess self-reported changes in food purchases and evaluate WLs impact was adapted from another IFPS measure previously used to evaluate the sugar-sweetened beverage tax policy [45]. However, results should be interpreted considering some limitations. The first is related to the non-probabilistic sampling design of the online survey, which prevents the extrapolation of the results to a national level. When comparing the characteristics of the sample in 2020 with those of the national census of 2020 [21, 22], it was observed that there were notably lower levels of overweight and obesity compared to national benchmark estimates. This fact underscores the importance of our subanalyses with the oversample of low educational level, which is more similar to the population of Mexican adults. Secondly, although this study assessed the frequency of beverage consumption, total calorie and critical nutrient intakes were not estimated; therefore, we were unable to evaluate whether overall dietary patterns were healthier among those who self-reported reducing their purchases of unhealthy products due to the WL. Furthermore, this study did not capture detailed information regarding the specific strategies employed to reduce the purchase of unhealthy foods. For instance, we were unable to determine whether the reduction was due to portion control, decreased frequency of consumption, or substitution with products that had fewer or no WLs.

In conclusion, our findings suggest that WLs contribute to a reduction in the unhealthy food and beverages purchases that pose risk to the health of the Mexican population and hold significant potential for promoting healthier decision-making among both Mexican youth and adults. Self-reported decreases in the purchases of unhealthy food products, especially sugary beverages, were more pronounced among subgroups characterized by lower education levels, a low-income adequacy, self-identified indigenous, and those with the highest intakes of water. These results underscore the positive impact of the labeling policy and highlight the opportunity to reinforce the promotion of healthier food choices within subgroups that self-reported fewer changes in their unhealthy food purchases. It is essential to maintain the implementation of this policy in the country and consistently promote its adoption.

## Abbreviations

IFPS: International Food Policy Study
WLs: warning labels
FoPL: front of package nutrition label
BMI: body mass index

## Supplementary Information

**Additional file 1.** Adjusted percentage of participants reporting that the warning labels had led them to buy less unhealthy foods overall and across food categories. IFPS 2020 and 2021

**Additional file 2.** Adjusted percentage of self-reported usefulness of each of the warning label in Mexican, youth and adults in 2020 and 2021.

**Additional file 3.** Odds ratio of perceiving each warning label most useful for food decision making in Mexican adults and youth in 2020 and 2021.

## Acknowledgements

We would like to thank nutritionist Celia Machorro for the literature search conducted for this study.

## Authors’ contributions

A.C. performed the statistical analysis, interpreted the data, and drafted the manuscript. C.N., K.Q. and J.V. helped draft the manuscript. A.C.M, A.J. and S.B. made substantial contributions in interpreting the data. J.T. provided statistical expertise and advice. CMW oversaw data collection and curation. D.H. conceived, designed, and executed the International Food Policy Study. All authors revised the manuscript critically for intellectual content. All authors read and approved the final manuscript.

## Availability of data and materials

The datasets used and/or analyzed during the current study are available from the corresponding author on reasonable request.

## Declarations

### Ethics approval and consent to participate

This study was conducted according to the guidelines laid down in the Declaration of Helsinki. The study was reviewed and received ethics clearance through a University of Waterloo Research Ethics Committee (ORE#41477 and #30829). Informed consent was obtained from all respondents prior to their participation.

A full description of the study methods can be found in the International Food Policy Study: Technical Reports (2020-2021) at www.foodpolicystudy.com/methods.

### Consent for publication

Not applicable

## Supplementary Information

**Additional file 2.** Adjusted percentage of participants reporting that the warning labels had led them to buy less unhealthy foods overall and across food categories. International Food Policy Study, 2020 and 2021.

Percentages were obtained from logistic regression models for each food group and adjusted by year of the survey, sex, age, indigeneity, income adequacy and BMI category for adults and youth, and additionally adjusted by educational level, children in the household, nutrition knowledge, role in the food shopping in the household for adults.

**Bold** numbers indicate significant difference between 2020 and 2021 (p<0.05)

**Additional Figure 1**. Odds ratio of perceiving buying less sugary drinks due to the WLs by quintile of the intake of sugary beverages and water in adults including an oversample with a low education level in 2021, International Food Policy Study, 2020 and 2021 (n=8,555).

Perceived purchase change in sugary drink groups included cola, soda, and sweetened fruit drinks. Sugary drink intake was based on the summed volume of regular soda, sweetened fruit drinks, regular flavored waters or vitamin waters with calories, regular sports drinks, and regular energy drinks. Median intake per quintile: Sugary drinks intake (Q1=71 ml, Q2=295 ml, Q3=526 ml, Q4=832 ml, Q5=1529 ml), Water intake (Q1=0 ml, Q2=357 ml, Q3=714 ml, Q4=1000 ml, Q5=1786 ml). Multilevel regression model adjusted by age, sex, indigeneity, educational level, income adequacy, children in the household, nutrition knowledge, role in the food shopping in the household, BMI category and year of the survey.

Additional file 3. Adjusted percentage of participants perceiving warning labels as useful in Mexican youth and adults, International Food Policy Study, 2020 and 2021.

Percentages were obtained from logistic regression models for each WLs and adjusted by year of the survey, sex, age, indigeneity, income adequacy and BMI category for youth and adults, and additionally adjusted by education level, children in the household, nutrition knowledge, and food shopping role in the household for adults.

Bold numbers indicate significant difference between 2020 and 2021 (p<0.05)

Additional file 4. Odds ratio of perceiving each warning label most useful for choosing healthier foods among Mexican adults and youth, International Food Policy Study, 2020 and 2021.

Odds ratios were obtained from logistic regression models.

