## Additional File 1 for "Self-reported decreases in the purchases of selected unhealthy foods resulting from the implementation of warning labels in Mexican youth and adult population"

| **Additional file 1**. Sociodemographic characteristics of the main sample of adults and oversample with a low-income adequacy. International Food Policy Study 2020 and 2021. | | | | | |
| --- | --- | --- | --- | --- | --- |
|  |  | **2020** | **2021**  **Main sample** | **2021**  **(Including the low-education level oversample)** | **Overall**  **(Including the low-education level oversample.)** |
| n sample |  | 3,900 | 3,875 | 5,497 | 9,397 |
|  |  | **% (95% CI)** | **% (95% CI)** | **% (95% CI)** | **% (95% CI)** |
| Age (years)* |  | 40.5 (39.9, 41.1) | 40.5 (40.3, 41.6) | 40.8 (40.2, 41.5) | 40.7 (40.2, 41.1) |
| Sex | Females | 51.8 (49.9, 53.7) | 52.0 (50.2, 53.8) | 51.9 (49.9, 53.8) | 51.9 (50.5, 53.2) |
|  | Males | 48.2 (46.3, 50.0) | 47.9 (46.1, 49.8) | 48.1 (46.2, 50.0) | 48.1 (46.8, 49.5) |
| Ethnicity | Yes | 19.2 (17.6, 20.9) | 19.2 (17.7, 20.9) | 19.4 (17.9, 20.9) | 19.3 (18.2, 20.4) |
|  | No | 80.8 (79.1, 82.4) | 80.8 (79.1, 82.3) | 80.6 (79.1, 82.1) | 80.7 (79.6, 81.8) |
| Income adequacy | Difficult | 50.2 (48.3, 52.1) | 39.8 (37.9, 41.7) | 47.9 (45.9, 49.8) | 48.8 (47.5, 50.2) |
|  | Neither | 36.1 (34.3, 37.9) | **40.8 (39.0, 42.7)** | 40.8 (38.9, 42.7) | 38.8 (37.5, 40.1) |
|  | Easy | 13.7 (12.6, 15.0) | **19.4 (18.0, 20.8)** | 11.4 (10.3, 12.5) | 12.4 (11.6, 13.2) |
| BMI category | <25 | 38.0 (36.2, 39.8) | 41.1 (39.3, 43.0) | 32.6 (30.9, 34.4)* | 34.9 (33.6, 36.2) |
|  | 25-29 | 31.9 (30.2, 33.8) | 30.6 (28.9, 32.3) | 29.5 (27.7, 31.2) | 30.5 (29.2, 31.8) |
|  | >=30 | 15.3 (13.9, 16.8) | 14.8 (13.5, 16.2) | 17.4 (15.9, 18.9) | 16.5 (15.5, 17.6) |
|  | Missing | 14.7 (13.4, 16.1) | **13.4 (12.1, 14.7)** | 20.5 (18.9, 22.2) | 18.1 (17.0, 19.2) |
| Education level | Low | 21.4 (19.9, 23.0) | 19.1 (17.7, 20.5) | 74.6 (73.0, 75.9)* | 52.3 (50.9, 53.7) |
|  | Medium | 13.6 (12.2, 14.9) | 14.2 (12.9, 15.6) | 10.0 (8.8, 11.4) | 11.5 (10.6, 12.5) |
|  | High | 65.0 (63.2, 66.8) | **66.8 (64.9, 68.5)** | 15.3 (14.5, 16.3) | 36.1 (34.9, 37.4) |
| Children in the household (<18 y old) | No | 51.2 (49.3, 53.1) | 50.4 (48.5, 52.3) | 48.8 (46.9, 50.7) | 49.7 (48.4, 51.1) |
|  | Yes | 48.7 (46.8, 50.6) | 49.6 (47.7, 51.4) | 51.3 (49.4 53.2) | 50.3 (48.9, 51.6) |
| Food shopping role (resume) | Important | 73.6 (72.0, 75.2) | 73.2 (71.6, 74.8) | 68.2 (66.4, 69.9)* | 70.5 (69.2, 71.7) |
|  | Some or None | 26.4 (24.7, 28.0) | 26.7 (25.1, 28.4) | 31.8 (30.0, 33.6) | 29.5 (28.3, 30.8) |
| Nutrition knowledge | Not knowledeable | 31.4 (29.7, 33.1) | 33.9 (32.2, 35.7) | 44.3 (42.4, 46.2)* | 38.9 (37.6, 40.2) |
|  | Somewhat | 55.5 (53.6, 57.4) | 50.8 (48.9, 52.7) | 46.3 (44.3, 48.2) | 50.1 (48.8, 51.5) |
|  | Knowledgeable | 13.1 (11.8, 14.5) | 15.2 (13.9, 16.6) | 9.4 (8.4, 10.6) | 10.9 (10.2, 11.8) |
| The mean and 95% CI are reported for age. **Bold numbers** indicate significant difference between 2020 and main sample in 2021 (p<0.05)  *Indicate significant difference between main sample in 2021 and including low-education level oversample. | | | | | |
