## Additional File 2 for "Self-reported decreases in the purchases of selected unhealthy foods resulting from the implementation of warning labels in Mexican youth and adult population"

| **Additional file 2.** Adjusted percentage of participants reporting that the warning labels had led them to buy less unhealthy foods overall and across food categories. International Food Policy Study, 2020 and 2021. | | | | |
| --- | --- | --- | --- | --- |
|  |  | **Youth** | **Adults** | **Adults** |
|  |  | **(n=1674)** | **(n=7,775)** | **(Including low-education level oversample) (n=9,397)** |
| Food group | **Year** | **% (95% CI)** | **% (95% CI)** | **% (95% CI)** |
| Overall (excluding 100% fruit juice) | 2020 | 39.5 (37.3, 41.7) | 45.4 (44.2, 46.6) | 47.5 (46.2, 48.7) |
|  | 2021 | 37.7 (35.6, 39.8) | 44.2 (43.1, 45.3) | 48.8 (47.7, 49.9) |
|  | Total | 38.6 (37.0, 40.3) | 44.8 (44.0, 45.6) | 48.3 (47.3, 49.1) |
| Cola | 2020 | 51.7 (47.7, 55.8) | 52.7 (50.7, 54.6) | 53.3 (51.1, 55.5) |
|  | 2021 | 47.9 (43.7, 52.0) | 52.1 (50.2, 54.0) | 55.4 (53.6, 57.3) |
|  | Total | 49.9 (47.0, 52.9) | 52.4 (51.0, 53.7) | 54.6 (53.1, 55.9) |
| Soda | 2020 | 51.3 (47.2, 55.3) | 53.0 (51.1, 54.9) | 53.9 (51.8, 56.1) |
|  | 2021 | 47.4 (43.1, 51.6) | 51.3 (49.4, 53.2) | 55.7 (53.8, 57.3) |
|  | Total | 49.4 (46.4, 52.4) | 52.2 (50.8, 53.5) | 54.9 (53.6, 56.3) |
| Diet soda | 2020 | 49.6 (45.5, 53.7) | 50.6 (48.7, 52.5) | 52.6 (50.5, 54.7) |
|  | 2021 | 45.0 (40.8, 49.3) | 48.8 (47.0, 50.7) | 53.5 (51.6, 55.3) |
|  | Total | 47.5 (44.4, 50.4) | 49.7 (48.4, 51.1) | 53.1 (51.7, 54.5) |
| Sweetened fruit drinks | 2020 | 42.5 (38.4, 46.6) | 50.6 (48.7, 52.5) | 51.3 (49.2, 53.4) |
|  | 2021 | 39.3 (35.2, 43.3) | 50.2 (48.3, 52.1) | 52.9 (51.1, 54.8) |
|  | Total | 41.0 (38.1, 43.9) | 50.4 (49.0, 51.7) | 52.2 (50.9, 53.7) |
| Candy or chocolate bars | 2020 | 39.9 (35.9, 43.9) | 46.3 (44.4, 48.3) | 47.6 (45.5, 49.8) |
|  | 2021 | 37.2 (33.2, 41.3) | 45.4 (43.6, 47.3) | **50.9 (49.1, 52.8)** |
|  | Total | 38.6 (35.8, 41.5) | 45.9 (44.5, 47.2) | 49.6 (48.2, 51.0) |
| Snacks such as chips | 2020 | 35.9 (32.0, 39.9) | 45.5 (43.6, 47.4) | 47.3 (45.2, 49.5) |
|  | 2021 | 34.2 (30.2, 38.2) | 42.9 (41.0, 44.7) | 46.9 (45.1, 48.7) |
|  | Total | 35.2 (32.4, 37.9) | 44.2 (42.8, 45.5) | 47.1 (45.7, 48.5) |
| Desserts such as cakes, | 2020 | 38.1 (34.1, 42.1) | 46.3 (44.4, 48.2) | 47.1 (44.9, 49.2) |
| cookies and ice cream | 2021 | 34.6 (30.5, 38.6) | 44.6 (42.7, 46.5) | 48.8 (46.9, 50.7) |
|  | Total | 36.4 (33.6, 39.2) | 45.4 (44.1, 46.8) | 48.1 (46.7, 49.5) |
| Sugary cereals | 2020 | 39.5 (35.5, 43.5) | 48.8 (46.9, 50.8) | 49.2 (47.0, 51.3) |
|  | 2021 | 38.0 (33.9, 42.1) | 49.4 (47.5, 51.2) | 51.9 (50.1, 53.8) |
|  | Total | 38.8 (35.9, 41.6) | 49.1 (47.8, 50.4) | 50.8 (49.4, 52.1) |
| 100% fruit or vegetable juice | 2020 | 21.0 (17.7, 24.4) | 29.6 (27.8, 31.3) | 29.2 (27.3, 31.2) |
|  | 2021 | 18.7 (15.4, 21.9) | **26.7 (25.0, 28.4)** | 27.4 (25.7, 29.1) |
|  | Total | 19.9 (17.6, 22.2) | 28.1 (26.9, 29.3) | 28.2 (26.9, 29.4) |
| Percentages were obtained from logistic regression models for each food group and adjusted by year of the survey, sex, age, indigeneity, income adequacy and BMI category for adults and youth, and additionally adjusted by educational level, children in the household, nutrition knowledge, role in the food shopping in the household for adults. Bold numbers indicate significant difference between 2020 and 2021 (p<0.05) | | | | |
