## Additional File 3 for "Self-reported decreases in the purchases of selected unhealthy foods resulting from the implementation of warning labels in Mexican youth and adult population"

| **Additional file 3.** Adjusted percentage of participants perceiving warning labels as useful in Mexican youth and adults, International Food Policy Study, 2020 and 2021. | | | | |
| --- | --- | --- | --- | --- |
|  |  | **Youth**  **(n=1,671)** | **Adults**  **(n=7,775)** | **Adults**  **(Including lo-education level oversample) (n=9,370)** |
|  | Year | **% (95% CI)** | **% (95% CI)** | **% (95% CI)** |
| Excess Calories | 2020 | 10.3 (7.8, 12.8) | 9.5 (8.4, 10.6) | 8.9 (7.8, 10.1) |
|  | 2021 | 10.8 (8.2, 13.3) | 10.2 (9, 11.5) | 10.6 (9.3, 11.8) |
|  | Total | 10.8 (9.0, 12.6) | 10.3 (9.5, 11.1) | 10.2 (9.3, 11.0) |
| Excess Sodium | 2020 | 3.4 (2.2, 4.7) | 6 (5, 6.9) | 5.5 (4.6, 6.5) |
|  | 2021 | 4.4 (2.4, 6.5) | 6.7 (5.8, 7.7) | 6.3 (5.5, 7.2) |
|  | Total | 4.3 (3.1, 5.4) | 6.6 (5.9, 7.3) | 6.2 (5.6, 6.9) |
| Excess Trans Fat | 2020 | 5.7 (3.8, 7.5) | 5.9 (5, 6.8) | 5.4 (4.4, 6.3) |
|  | 2021 | 5.7 (3.6, 7.9) | 6.3 (5.4, 7.2) | 5.7 (4.9, 6.6) |
|  | Total | 6.0 (4.6, 7.5) | 6.3 (5.7, 7) | 5.8 (5.2, 6.4) |
| Excess Sugars | 2020 | 22.8 (19.4, 26.2) | 16.8 (15.3, 18.2) | 17.8 (16.2, 19.5) |
|  | 2021 | 23.3 (19.9, 26.8) | 18.8 (17.3, 20.2) | 18.1 (16.7, 19.5) |
|  | Total | 23.1 (20.7, 25.6) | 18.2 (17.2, 19.2) | 18.3 (17.3, 19.4) |
| Excess Saturated Fat | 2020 | 8.4 (6.2, 10.6) | 9 (7.9, 10) | 9.2 (7.9, 10.5) |
|  | 2021 | 9.1 (6.6, 11.7) | 10.1 (9, 11.2) | **11.3 (10.2, 12.5)** |
|  | Total | 9.1 (7.3, 10.8) | 9.7 (8.9, 10.5) | 10.6 (9.7, 11.4) |
| None have been useful | 2020 | 15.2 (12.3, 18.2) | 16.2 (14.8, 17.6) | 16.4 (14.8, 18.0) |
|  | 2021 | 14.9 (11.9, 18.1) | 15.4 (14.1, 16.8) | 142. (12.9, 15.5) |
|  | Total | 15.3 (13.2, 17.4) | 16.7 (15.7, 17.7) | 15.8 (14.8, 16.8) |
| All have been equally useful | 2020 | 32.4 (28.5, 36.4) | 34.1 (32.2, 35.9) | 34.6 (32.5, 36.6) |
|  | 2021 | 29.6 (25.9, 33.4) | **29.6 (27.9, 31.3)** | **31.4 (29.7, 33.2)** |
|  | Total | 31.3 (28.6, 33.9) | 32.1 (30.9, 33.4) | 33.0 (31.7, 34.3) |
| Percentages were obtained from logistic regression models for each WLs and adjusted by year of the survey, sex, age, indigeneity, income adequacy and BMI category for youth and adults, and additionally adjusted by education level, children in the household, nutrition knowledge, and food shopping role in the household for adults.  **Bold numbers** indicate significant difference between 2020 and 2021 (p<0.05) | | | | |
