## Additional File 4 for "Self-reported decreases in the purchases of selected unhealthy foods resulting from the implementation of warning labels in Mexican youth and adult population"

| Additional file 4. Odds ratio of perceiving each warning label most useful for choosing healthier foods among Mexican adults and youth, International Food Policy Study, 2020 and 2021. | | | | | | | | |
| --- | --- | --- | --- | --- | --- | --- | --- | --- |
| **Adults (n=7,752)** | WL | Calories | Sodium | Trans Fat | Sugars | Saturated fat | None | All |
| Category | Subpopulation | OR (95% CI) | OR (95% CI) | OR (95% CI) | OR (95% CI) | OR (95% CI) | OR (95% CI) | OR (95% CI) |
| Age | Years | **0.98 (0.98, 0.99)** | 1 (0.99, 1.01) | 1 (0.99, 1) | **0.99 (0.98, 0.99)** | 1 (0.99, 1) | **1.01 (1, 1.01)** | **1.01 (1.01, 1.02)** |
| Sex | Female | 1 | 1 | 1 | 1 | 1 | 1 | 1 |
|  | Male | 0.95 (0.79, 1.14) | **0.76 (0.61, 0.94)** | 0.79 (0.62, 1) | **1.28 (1.11, 1.48)** | 0.98 (0.82, 1.17) | **1.19 (1.03, 1.38)** | 0.9 (0.8, 1.01) |
| Indigeneity | No | 1 | 1 | 1 | 1 | 1 | 1 | 1 |
|  | Si | **1.4 (1.11, 1.76)** | 0.97 (0.71, 1.34) | 1.2 (0.87, 1.65) | **1.23 (1.03, 1.47)** | 1.12 (0.87, 1.44) | **0.56 (0.45, 0.7)** | 0.93 (0.79, 1.1) |
| Educational level | High | 1 | 1 | 1 | 1 | 1 | 1 | 1 |
|  | Medium | **0.71 (0.54, 0.94)** | 0.95 (0.67, 1.34) | 1.14 (0.82, 1.58) | 0.94 (0.76, 1.17) | 0.91 (0.68, 1.21) | 0.98 (0.79, 1.22) | **1.22 (1.02, 1.45)** |
|  | Low | 0.84 (0.66, 1.06) | 0.78 (0.59, 1.02) | 0.75 (0.55, 1.03) | 1.11 (0.93, 1.32) | 1.11 (0.89, 1.4) | 0.98 (0.81, 1.18) | 1.12 (0.96, 1.3) |
| Income adequacy | Difficult | **0.69 (0.54, 0.88)** | 1.35 (0.99, 1.86) | 1.13 (0.83, 1.55) | 0.91 (0.75, 1.1) | 0.85 (0.66, 1.09) | 1.16 (0.93, 1.44) | 1.13 (0.95, 1.34) |
|  | Neither | **0.76 (0.6, 0.96)** | **1.48 (1.09, 2.01)** | 1.07 (0.79, 1.45) | 0.9 (0.74, 1.08) | 0.85 (0.67, 1.08) | 1.03 (0.82, 1.28) | 1.16 (0.98, 1.38) |
|  | Easy | 1 | 1 | 1 | 1 | 1 | 1 | 1 |
| Children in the household | No | 1 | 1 | 1 | 1 | 1 | 1 | 1 |
|  | Yes | 1.06 (0.88, 1.28) | **1.33 (1.05, 1.67)** | 1.04 (0.83, 1.31) | **1.32 (1.15, 1.53)** | 1.1 (0.92, 1.33) | **0.76 (0.66, 0.88)** | **0.84 (0.75, 0.95)** |
| Nutritional knowledge | Little | 1 | 1 | 1 | 1 | 1 | 1 | 1 |
|  | Somewhat know | 1.07 (0.88, 1.3) | 0.89 (0.7, 1.13) | 1.29 (0.99, 1.69) | 1.01 (0.87, 1.17) | 1.14 (0.93, 1.4) | **0.57 (0.49, 0.67)** | **1.28 (1.12, 1.46)** |
|  | Knowledgeable | 1.21 (0.92, 1.58) | 1.16 (0.81, 1.67) | 1.31 (0.9, 1.91) | 1.05 (0.84, 1.31) | 0.87 (0.63, 1.2) | **0.51 (0.4, 0.65)** | **1.29 (1.06, 1.56)** |
| Shopping role | No | 1 | 1 | 1 | 1 | 1 | 1 | 1 |
|  | Yes | 0.87 (0.71, 1.07) | 1.01 (0.79, 1.29) | 1.03 (0.76, 1.41) | 1.14 (0.96, 1.35) | 1.13 (0.92, 1.39) | 0.97 (0.83, 1.14) | 0.93 (0.81, 1.06) |
| BMI category | Normal | 1 | 1 | 1 | 1 | 1 | 1 | 1 |
|  | Overweight | 1.2 (0.96, 1.49) | 0.91 (0.7, 1.18) | 1.14 (0.87, 1.49) | 0.95 (0.8, 1.13) | 0.84 (0.68, 1.04) | 1.14 (0.95, 1.35) | 0.95 (0.82, 1.09) |
|  | Obesity | 1.16 (0.88, 1.53) | 1.08 (0.78, 1.49) | **0.66 (0.45, 0.97)** | 1.09 (0.88, 1.35) | **0.71 (0.53, 0.96)** | **1.25 (1.02, 1.55)** | 0.94 (0.78, 1.13) |
|  | Missing | 1.23 (0.95, 1.6) | **0.61 (0.42, 0.87)** | 1.23 (0.88, 1.73) | 1.17 (0.95, 1.45) | 0.91 (0.68, 1.2) | 1 (0.79, 1.26) | 0.9 (0.74, 1.09) |
| Year | 2020 | 1 | 1 | 1 | 1 | 1 | 1 | 1 |
|  | 2021 | 1.08 (0.91, 1.3) | 1.14 (0.91, 1.42) | 1.07 (0.85, 1.34) | 1.15 (1, 1.32) | 1.14 (0.96, 1.37) | 0.95 (0.82, 1.09) | **0.81 (0.72, 0.92)** |
| **Adults** (main+oversample)  **(n=9,370)** | WL | Calories | Sodium | Trans Fat | Sugars | Saturated fat | None | All |
| Category | Subpopulation | OR (95% CI) | OR (95% CI) | OR (95% CI) | OR (95% CI) | OR (95% CI) | OR (95% CI) | OR (95% CI) |
| Age | Years | **0.99 (0.98, 1)** | 1 (0.99, 1.01) | 1 (0.99, 1.01) | **1.0 (0.99, 1)** | **1 (0.99, 1)** | **1.01 (1.01, 1.02)** | **1.02 (1.01, 1.02)** |
| Sex | Female | 1 | 1 | 1 | 1 | 1 | 1 | 1 |
|  | Male | 0.96 (0.8, 1.16) | **0.79 (0.63, 0.99)** | 1 (0.79, 1.27) | **1.33 (1.15, 1.54)** | 1.02 (0.85, 1.22) | 1.14 (0.98, 1.33) | **0.83 (0.74, 0.94)** |
| Indigeneity | No | 1 | 1 | 1 | 1 | 1 | 1 | 1 |
|  | Yes | **1.33 (1.06, 1.65)** | 1.02 (0.76, 1.38) | 1.23 (0.91, 1.65) | 1.09 (0.92, 1.31) | 1.08 (0.85, 1.36) | **0.65 (0.52, 0.8)** | 0.98 (0.83, 1.14) |
| Educational level | High | 1 | 1 | 1 | 1 | 1 | 1 | 1 |
|  | Medium | 0.95 (0.69, 1.32) | 0.92 (0.62, 1.35) | 1.1 (0.76, 1.61) | 0.82 (0.64, 1.04) | 1.13 (0.84, 1.52) | 0.89 (0.7, 1.13) | 1.18 (0.97, 1.44) |
|  | Low | **0.82 (0.68, 1)** | **0.79 (0.61, 1.01)** | 0.79 (0.6, 1.04) | **1.2 (1.02, 1.4)** | 1.16 (0.95, 1.42) | **0.83 (0.7, 0.99)** | 1.15 (1.01, 1.32) |
| Income adequacy | Easy | 1 | 1 | 1 | 1 | 1 | 1 | 1 |
|  | Neither | 0.89 (0.69, 1.16) | 1.33 (0.94, 1.88) | 1.00 (0.70, 1.41) | 0.94 (0.76, 1.16) | 0.84 (0.65, 1.10) | 0.97 (0.76, 1.24) | 1.12 (0.92, 1.34) |
|  | Difficult | 0.77 (0.59, 1.00) | 1.37 (0.96, 1.93) | 1.22 (0.86, 1.72) | 0.94 (0.77, 1.15) | 0.89 (0.68, 1.16) | 1.04 (0.81, 1.32) | 1.05 (0.87, 1.26) |
| Children in the household | No | 1 | 1 | 1 | 1 | 1 | 1 | 1 |
|  | Yes | 0.98 (0.81, 1.18) | **1.33 (1.05, 1.67)** | 1.2 (0.95, 1.53) | **1.27 (1.1, 1.47)** | 1.05 (0.87, 1.26) | 0.89 (0.76, 1.04) | **0.82 (0.73, 0.93)** |
| Nutritional knowledge | Little | 1 | 1 | 1 | 1 | 1 | 1 | 1 |
|  | Somewhat know | 1.19 (0.97, 1.46) | 0.81 (0.64, 1.04) | **1.45 (1.1, 1.9)** | 0.96 (0.82, 1.11) | 1.16 (0.95, 1.42) | **0.58 (0.49, 0.68)** | **1.24 (1.09, 1.41)** |
|  | Knowledgeable | 1.18 (0.87, 1.59) | 1.01 (0.68, 1.48) | 1.32 (0.88, 1.96) | 1.07 (0.83, 1.36) | 1.01 (0.73, 1.41) | **0.49 (0.38, 0.63)** | **1.29 (1.05, 1.59)** |
| Shopping role | Some or none | 1 | 1 | 1 | 1 | 1 | 1 | 1 |
|  | Most | 1.07 (0.87, 1.33) | 0.99 (0.77, 1.29) | 1.07 (0.81, 1.42) | 1.09 (0.92, 1.3) | 1.15 (0.94, 1.42) | 0.89 (0.75, 1.06) | 0.93 (0.81, 1.06) |
| BMI category | Normal | 1 | 1 | 1 | 1 | 1 | 1 | 1 |
|  | Overweight | 1.06 (0.84, 1.35) | 0.96 (0.73, 1.26) | 1.11 (0.84, 1.48) | 1 (0.84, 1.2) | 1.03 (0.83, 1.28) | 1 (0.83, 1.2) | 0.97 (0.83, 1.12) |
|  | Obesity | 1.22 (0.92, 1.62) | 0.98 (0.7, 1.36) | 0.68 (0.47, 1.01) | 1.15 (0.93, 1.43) | 0.89 (0.67, 1.19) | 1.2 (0.96, 1.49) | 0.87 (0.73, 1.05) |
|  | Missing | 1.22 (0.94, 1.59) | **0.61 (0.43, 0.87)** | 1.05 (0.74, 1.5) | 1.05 (0.85, 1.29) | 1.05 (0.81, 1.36) | 1.21 (0.97, 1.52) | 0.88 (0.73, 1.05) |
| Year | 2020 | 1 | 1 | 1 | 1 | 1 | 1 | 1 |
|  | 2021 | 1.21 (1, 1.45) | 1.15 (0.92, 1.45) | 1.08 (0.85, 1.38) | 1.02 (0.88, 1.19) | **1.26 (1.04, 1.52)** | **0.85 (0.73, 0.99)** | **0.87 (0.77, 0.99)** |
| Youth (n=1,671) | WL | Calories | Sodium | Trans Fat | Sugars | Saturated fat | None | All |
| Category | Subpopulation | OR (95% CI) | OR (95% CI) | OR (95% CI) | OR (95% CI) | OR (95% CI) | OR (95% CI) | OR (95% CI) |
| Age | Years | 0.94 (0.79, 1.13) | 0.98 (0.77, 1.23) | 1.04 (0.83, 1.29) | 0.98 (0.87, 1.11) | 1.01 (0.83, 1.22) | 1.09 (0.94, 1.26) | 1 (0.89, 1.12) |
| Sex | Female | 1 | 1 | 1 | 1 | 1 | 1 | 1 |
|  | Male | 0.77 (0.52, 1.13) | 1.12 (0.62, 1.99) | 0.82 (0.49, 1.37) | 1.06 (0.8, 1.4) | 1.01 (0.67, 1.54) | 0.98 (0.7, 1.36) | 1.14 (0.89, 1.47) |
| Indigeneity | No | 1 | 1 | 1 | 1 | 1 | 1 | 1 |
|  | Yes | 1.03 (0.59, 1.79) | 0.51 (0.16, 1.61) | 1.75 (0.91, 3.39) | 1.21 (0.81, 1.79) | 1.55 (0.92, 2.6) | 0.81 (0.48, 1.37) | 0.76 (0.52, 1.1) |
| Income adequacy | Easy | 1 | 1 | 1 | 1 | 1 | 1 | 1 |
|  | Neither | 0.65 (0.41, 1.03) | 0.78 (0.41, 1.49) | 1.23 (0.66, 2.28) | 1.06 (0.73, 1.54) | 0.84 (0.49, 1.43) | 1.19 (0.74, 1.91) | 1.14 (0.82, 1.58) |
|  | Difficult | **0.59 (0.37, 0.96)** | **0.46 (0.22, 0.97)** | 0.74 (0.36, 1.52) | 0.90 (0.61, 1.34) | 0.96 (0.54, 1.70) | 1.52 (0.94, 2.45) | 1.37 (0.98, 1.90) |
| BMI category | Normal | 1 | 1 | 1 | 1 | 1 | 1 | 1 |
|  | Overweight | 1.34 (0.83, 2.16) | 0.84 (0.43, 1.65) | 1.59 (0.83, 3.07) | 0.93 (0.66, 1.31) | 0.66 (0.36, 1.2) | 1.2 (0.78, 1.84) | 0.87 (0.63, 1.19) |
|  | Obesity | 1.18 (0.52, 2.7) | 1.15 (0.33, 4.03) | 0.84 (0.24, 2.95) | 0.75 (0.38, 1.52) | 1.29 (0.54, 3.06) | 1.21 (0.6, 2.42) | 0.94 (0.53, 1.66) |
|  | Missing | 0.96 (0.58, 1.57) | 0.84 (0.4, 1.73) | 1.15 (0.59, 2.25) | 1.03 (0.71, 1.47) | 1.26 (0.73, 2.16) | 1.07 (0.69, 1.67) | 0.88 (0.63, 1.24) |
| Year | 2020 | 1 | 1 | 1 | 1 | 1 | 1 | 1 |
|  | 2021 | 1.06 (0.73, 1.52) | 1.31 (0.77, 2.22) | 1.02 (0.61, 1.72) | 1.04 (0.79, 1.37) | 1.1 (0.72, 1.7) | 0.98 (0.71, 1.36) | 0.88 (0.69, 1.13) |
| Odds ratios were obtained from logistic regression models. | | | | | | | | |
