## Supplementary figures and images for "Self-reported decreases in the purchases of selected unhealthy foods resulting from the implementation of warning labels in Mexican youth and adult population"

### Additional Figure 1

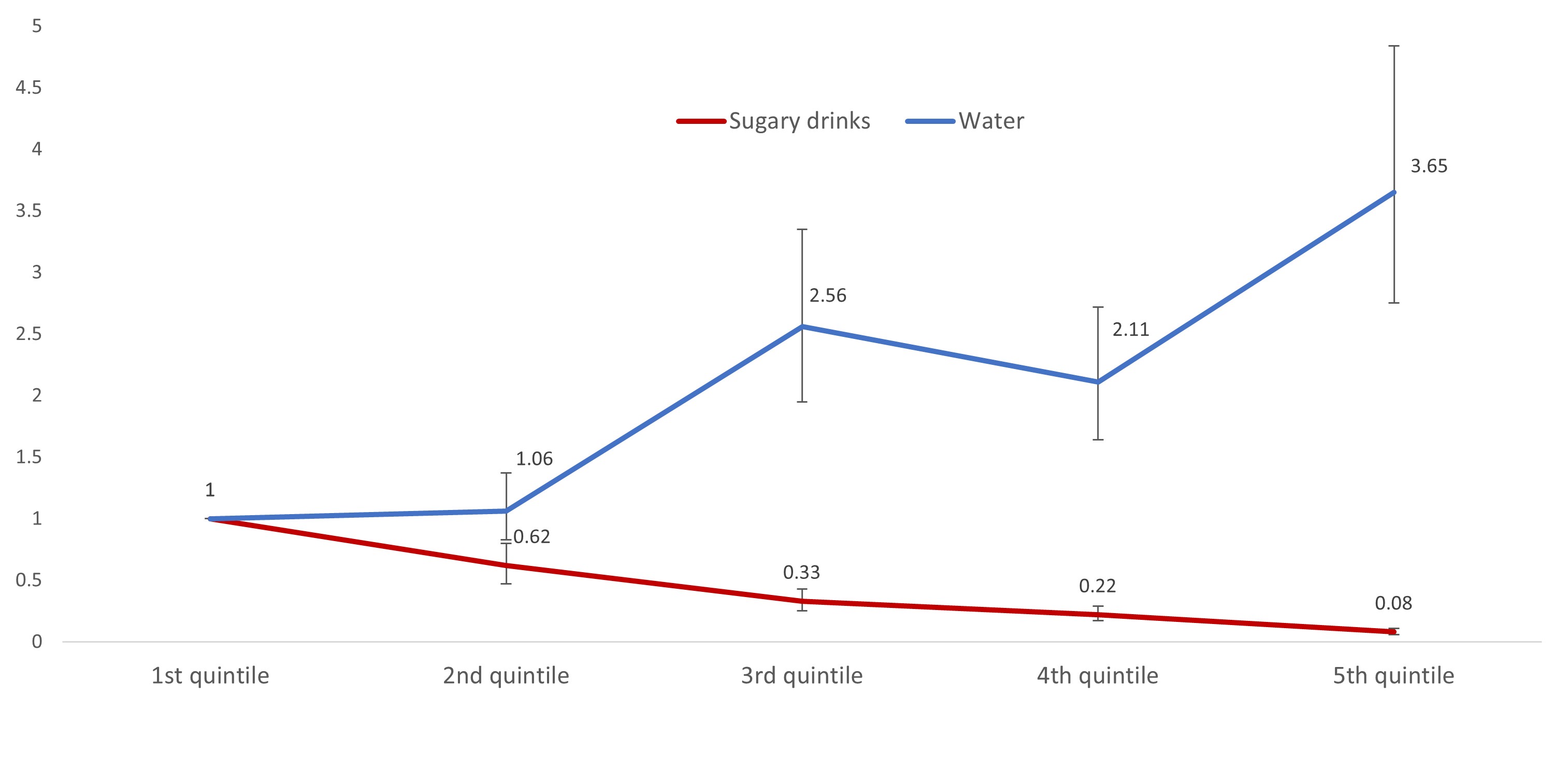
